# Adults with Severe Acute Respiratory Illness in Glasgow, 2021-22: A Prospective Cohort Study

**DOI:** 10.1101/2024.08.08.24311641

**Authors:** Antonia Ho, Neil McInnes, Andrew Blunsum, Joanna Quinn, Daniel Lynagh, Michael E Murphy, Rory Gunson, Alisdair MacConnachie, David J Lowe

## Abstract

**Objectives:** We report the findings of a novel enhanced syndromic surveillance which characterised influenza- and SARS-CoV-2-associated severe acute respiratory illness (SARI) in the 2021/2022 winter season.

**Methods:** Prospective cohort study of adults admitted to the Queen Elizabeth University Hospital, Glasgow, with a severe acute respiratory illness. Patient demographics, clinical history, admission details, and outcome were recorded.

**Results:** Between November 2021 and May 2022, 1,063 hospitalised SARI episodes in 1,037 adult patients were identified. Median age was 72.0 years and 44.5% were male. Most (82.6%) SARI cases had ≥1 co-morbidity; chronic lung disease (50.0%) and malignancy (22.5%) were the most frequently reported.

Overall, 229 (22%) and 33 (3%) SARI episodes were SARS-CoV-2 and influenza A PCR positive, respectively. Overall, 74.7%, 6.5% and 43.0% SARI episodes received antibiotics, antivirals, and steroids, respectively (54.5, 11.0 and 51.3% among COVID-19 patients). 1.1% required mechanical ventilation and 7.8% died. Male sex, multimorbidity, frailty, respiratory rate >30, low GCS and chest X-ray consolidation were predictive of in-hospital mortality.

**Conclusion:** Our syndromic surveillance provided near real-time data on hospitalised SARI to clinicians and Public Health Scotland, and informed them of the evolving clinical epidemiology of SARS-CoV-2 and influenza as we transition from the pandemic phase.

## Background

Before the emergence of severe acute respiratory syndrome coronavirus 2 (SARS-CoV-2), respiratory viral surveillance in the UK predominantly focused on influenza viruses.^1^ The impact of influenza on secondary care was monitored through laboratory-confirmed influenza cases admitted to hospital and critical care (high dependency units (HDU) and intensive care units (ICU)).^1^

While 2020 and 2021 were dominated by multiple waves of SARS-CoV-2 infections, influenza case numbers were low.^2^ Only 0.2% of respiratory samples in 2020-21 tested positive for influenza, compared to >25% positivity in the two preceding years.^2^ This was likely due to a combination of the implementation of non-pharmacological interventions (NPIs) to mitigate COVID-19 in the community as well as higher vaccine uptake in existing risk groups and extension of the flu vaccine programme to include 50-64 years old.^2^

Anticipated co-circulation of influenza viruses and SARS-CoV-2 prompted guidance from the World Health Organization (WHO)^3^ and the European Centre for Disease Prevention and Control (ECDC)^4^ for pathogen-agnostic syndromic surveillance of hospitalised patients with severe acute respiratory illness (SARI). Combined laboratory and clinical data, including reported symptoms, are needed to understand the evolving prevalence, clinical presentation, risk factors and outcomes of hospitalised influenza and SARS-CoV-2 infections. Moreover, timely data will inform clinical management, infection control, service planning, and public health intelligence, thereby improving clinical care delivery. Lastly, a platform to monitor for the ever-present threat of emerging and re-emerging respiratory pathogens is essential.

We described the implementation of a novel enhanced syndromic surveillance in a large tertiary hospital in Glasgow and the epidemiology of hospitalised SARI in the 2021/2022 winter season.

## Methods

### Setting and study population

The study, funded by Public Health Scotland (PHS), was conducted at the Queen Elizabeth University Hospital (QEUH), one of two large tertiary teaching adult hospitals in Glasgow that serve a population of 1.3 million. We performed a prospective cohort study of adults (aged ≥16 years) admitted to the QEUH with SARI, defined as: i) an individual presenting with an acute respiratory illness, such as fever, cough, dyspnoea, sore throat and malaise; ii) that required testing for influenza ± SARS-CoV-2 in the opinion of the treating clinician; and iii) required hospital admission. Full eligibility criteria are detailed in Figure 2.

**Figure 1.**
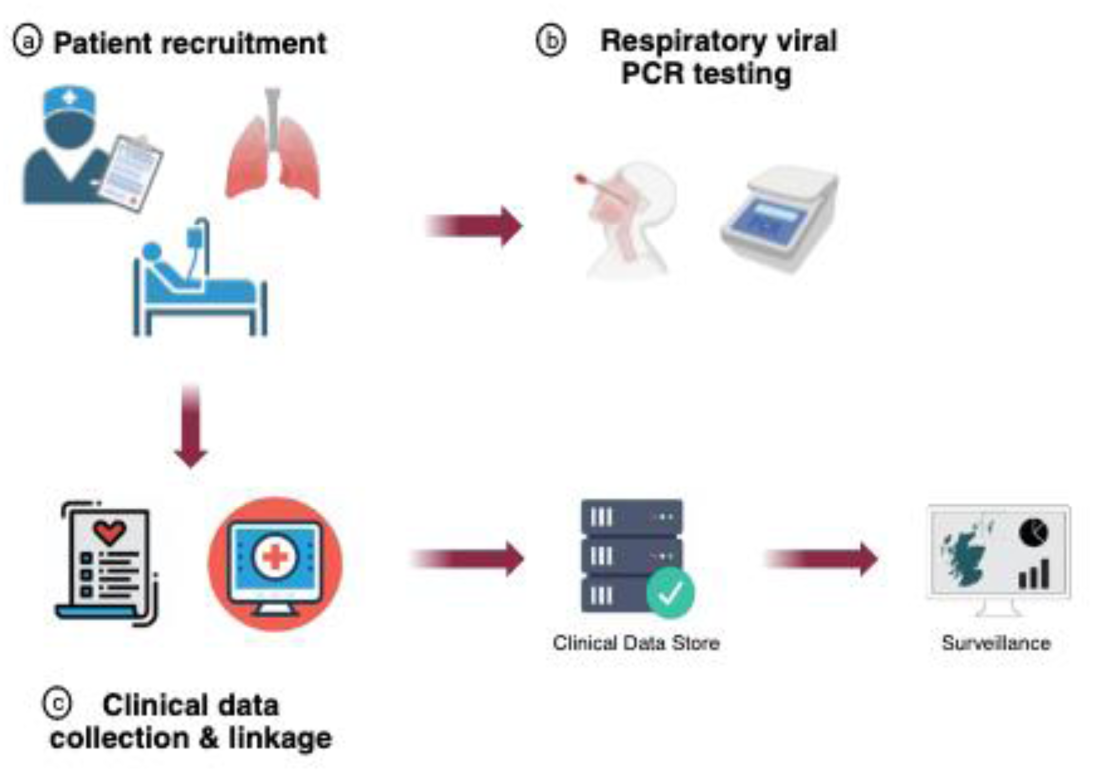
Study procedures. Figure made with Biorender.

**Figure 2.**
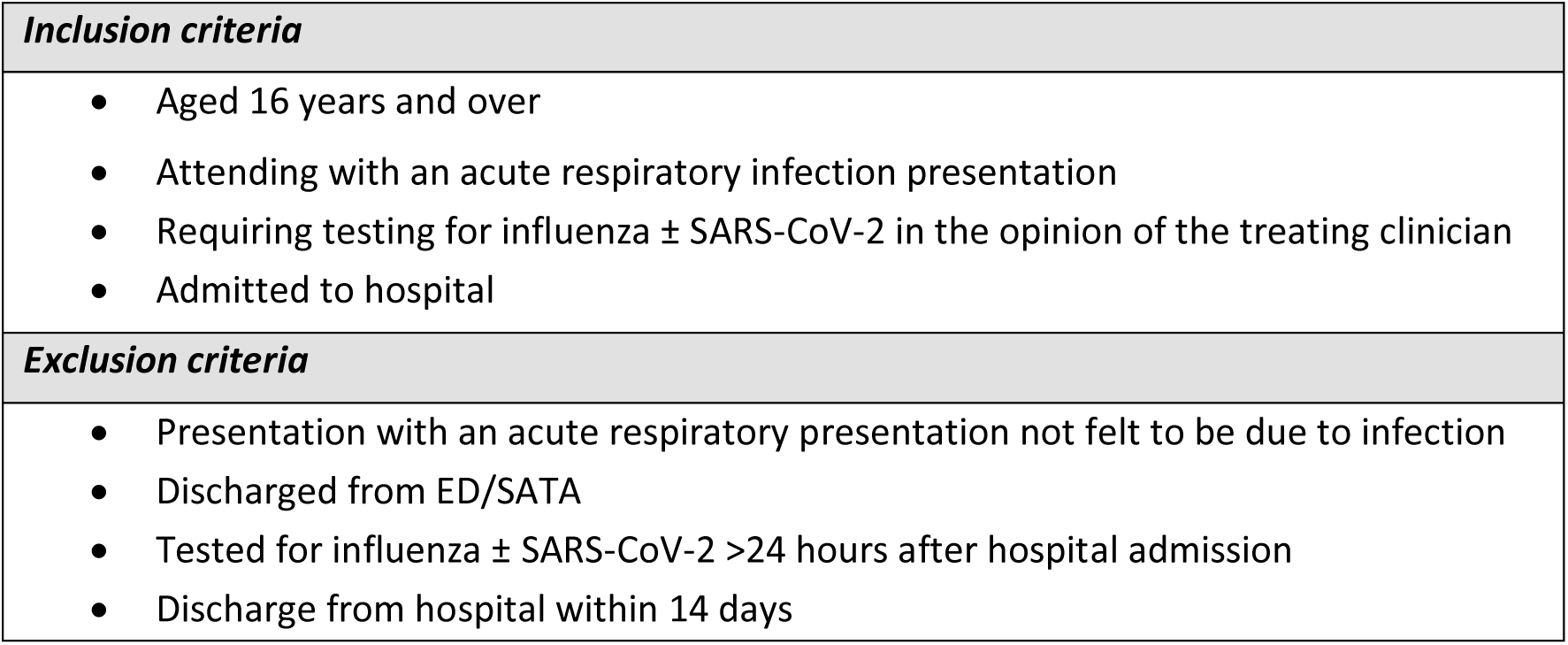
Study eligibility criteria. ED, Emergency Department; SATA, Specialist Assessment and Treatment Area.

Adult patients attending the Emergency Department (ED), and the Specialist Assessment and Treatment Area (SATA), the latter a dedicated assessment area for acute respiratory or unspecified febrile illness referrals from general practitioners (GPs), were screened for SARI by a dedicated research team. Recruitment took place between 23 October 2021 and 22 May 2022.

As routine clinical care, combined nose and throat swabs were tested for SARS-CoV-2 in addition to influenza A and B by real-time polymerase chain reaction (RT-PCR) using the point-of-care Cobas^®^ Liat^®^ system. Specimens PCR-positive for either virus were sent to the West of Scotland Specialist Virology Centre for confirmatory laboratory-based PCR and sequencing.

### Data collection, linkage and reporting

For patients that fulfilled eligbility criteria, data were collected and stored on the NHS Education for Scotland Digital (NES Digital) Turas Clinical Assessment Tool (TCAT), which was designed during the COVID-19 pandemic to collect detailed data on COVID-19 cases in various clinical settings and adapted for this study. On admission, data were collected on age, sex, healthcare worker status, presenting symptoms, Rockwood Clinical Frailty Scale (score ≥5 indicated frailty), influenza/COVID-19 vaccination status, admission physiological parameters, and influenza A/B and SARS-CoV-2 POC PCR result. Initial diagnosis and treatment given in ED/SATA were also recorded.

On discharge, the research team recorded additional details on the patient journey using electronic case records (Trakcare, Clinical Portal (Orion Health), Hospital Electronic Prescribing and Medicines Administration (HEPMA), and Scottish Care Information (SCI) gateway), including antimicrobials, antivirals and steroids prescribed during admission, maximum level of care (including HDU/ICU admission) and respiratory support, in addition to discharge date and patient disposition.

Additional patient data were obtained through linkage using the Community Health Index (CHI; a unique identifier used in all healthcare contacts across NHS Scotland) to electronic health records on socioeconomic status, pre-existing health conditions, laboratory and radiology results, SARS-CoV-2 variant, hospital journey, discharge diagnoses, and death registry (Supplementary Figure 1). Cohorts and de-identified linked data were prepared by the West of Scotland Safe Haven at National Health Service Greater Glasgow and Clyde (NHSGGC). An anonymised extract of the TCAT database was transferred to PHS weekly via NHSGGC Safe Haven in accordance with the study data sharing agreement. As per conventions for statistical disclosure control,^5^ the threshold for privacy risk of ‘n’ observations have been set to 5. Value less than 5 has been ascribed ‘<5’ in the text, tables and figures.

### Outcomes

The primary outcome was the weekly proportion of hospitalised SARI PCR-positive for SARS-CoV-2 or influenza A and B. Additionally, we characterised the clinical phenotype of patients admitted to hospital with influenza- and SARS-CoV-2-assocated SARI, including clinical presentation, circulating variants (for SARS-CoV-2), the prevalence and aetiology of significant bacterial infection (defined as a clinically relevant organism identified from respiratory (including sputum, respiratory aspirate and pleural fluid) or blood culture, or a positive urinary legionella antigen collected between 7 days before and 5 days post-admission, clinical severity (proportion of patients requiring respiratory support or critical care admission, and in-hospital death), and factors associated with in-hospital death. Lastly, the discriminatory power of two prognostic tools: i) 4C mortality score^6^ (validated in hospitalised COVID-19 patients)^6^; and ii) CURB-65^7^ (validated in community-acquired pneumonia) in identifying patients at risk of in-hospital death among hospitalised adult SARI patients with and without COVID-19 was evaluated.

### Statistical analysis

Patient characteristics were descriptively summarised. Categorical data were presented as counts and percentages, while continuous data were presented as either means with standard deviations (SD) or medians with inter-quartile ranges (IQR). We compared clinical presentation, frequency and aetiology of bacterial infections, antimicrobial use, patient journey, and outcome by SARS-CoV-2 PCR status. Categorical variables were compared using Chi-squared or Fisher’s exact test. Continuous variables were compared using Wilcoxon rank sum or Student’s t-test.

Predictors of in-hospital mortality were identified using backward stepwise logistic regression. Potential predictors evaluated include patient demographics, virological factors, as well as admission clinical parameters and investigation results. Adjusted odds ratios (aOR) and 95% confidence intervals (CI) were reported. We assessed the discrimination of 4C mortality score and CURB-65 using the area under the receiver operating characteristic (AUROC) curve, Overall goodness of fit was assessed using the Brier score (range 0 to 1); smaller values indicated better model performance. Calibration was assessed using calibration slopes, calibration in-the-large (CITL) and calibration plots. Data analyses were performed using Stata (v18.0).

### Ethics and permission

Ethics approval was obtained from the North of Scotland Research Ethics Committee (ref. 21/NS/1036) and the NHSGGC Biorepository (ref. 16/WS/0207) to store residual blood and respiratory samples. In Scotland, patient consent is not required when routinely collected patient data are used for research purposes through an approved Safe Haven. For that reason, informed consent was not required and was not sought.

## Results

### Cohort characteristics

Overall, 1063 SARI episodes from 1037 individuals were included between 23 November 2021 and 22 May 2022 (Supplementary Figure 2). SARI case numbers ranged from 20 in week 48 in 2021 to 65 in week 1 in 2022 (Figure 3). Twenty-six individuals had two or more episodes, and 461 (44.5%) of SARI cases were male (Table 1). Median age was 72.0 years (interquartile range (IQR) 60.1-81.6 years); nearly two-thirds (65.1%) were aged 65 years and over. The majority of recruited SARI cases (85.2%, n=906) were white, while 5.0% (n=53) were South Asian. Most (82.6%, n=857) had at least one pre-existing health condition, most commonly chronic lung disease (50.0%, n=518), followed by malignancy (22.5%, n=233) and diabetes (21.7%, n=225). Over half of recruited SARI cases (57.3%) had a CCI score of ≥5. Additionally, nearly 40% of the cohort were from the most deprived quintile in

**Figure 3.**
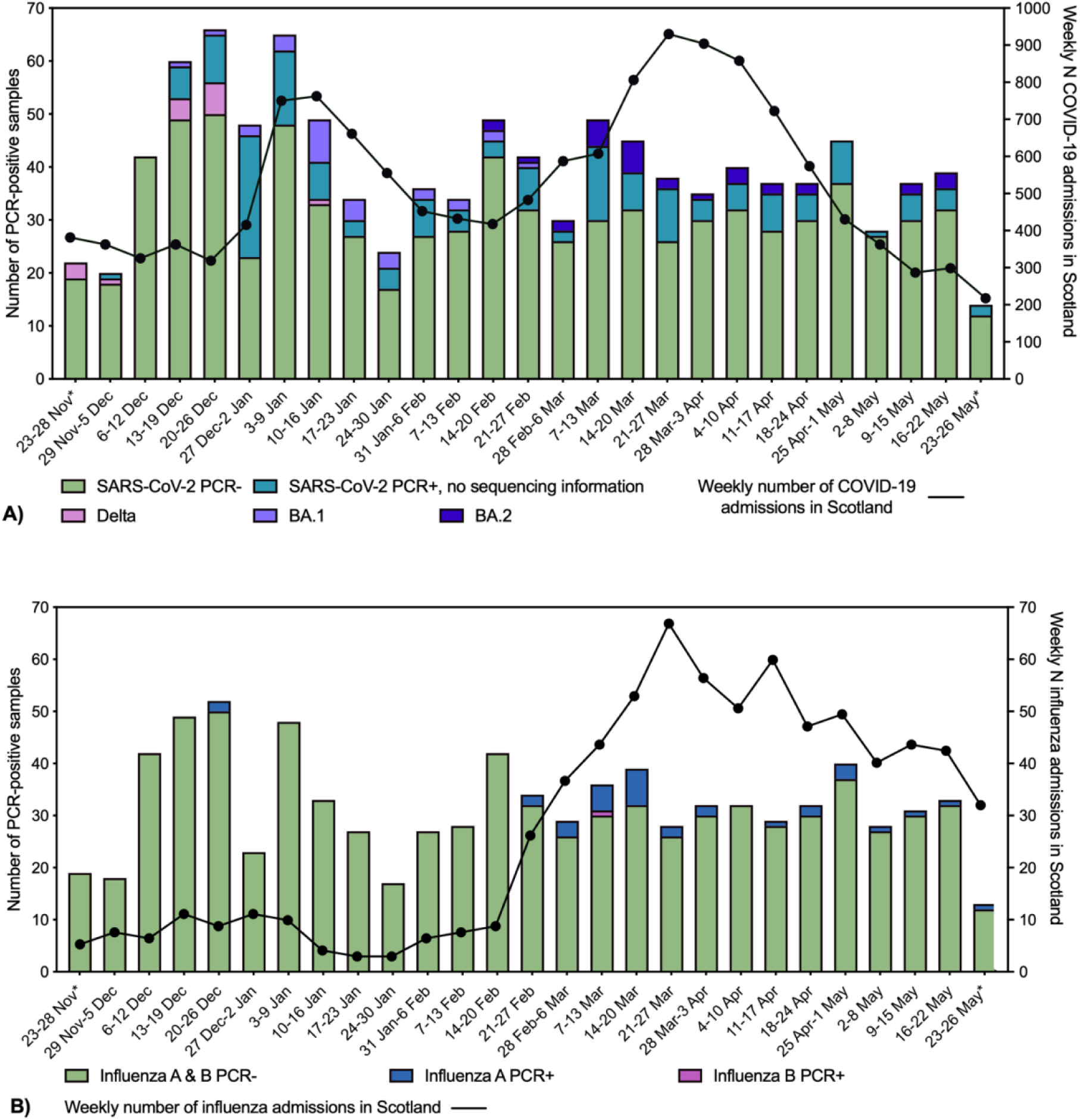
Number of SARI cases recruited weekly, with proportion that were **A)** SARS-CoV-2 PCR positive, including sequencing data where available; B) Influenza A & B PCR positive. *Incomplete week of recruitment.

**Table 1.**
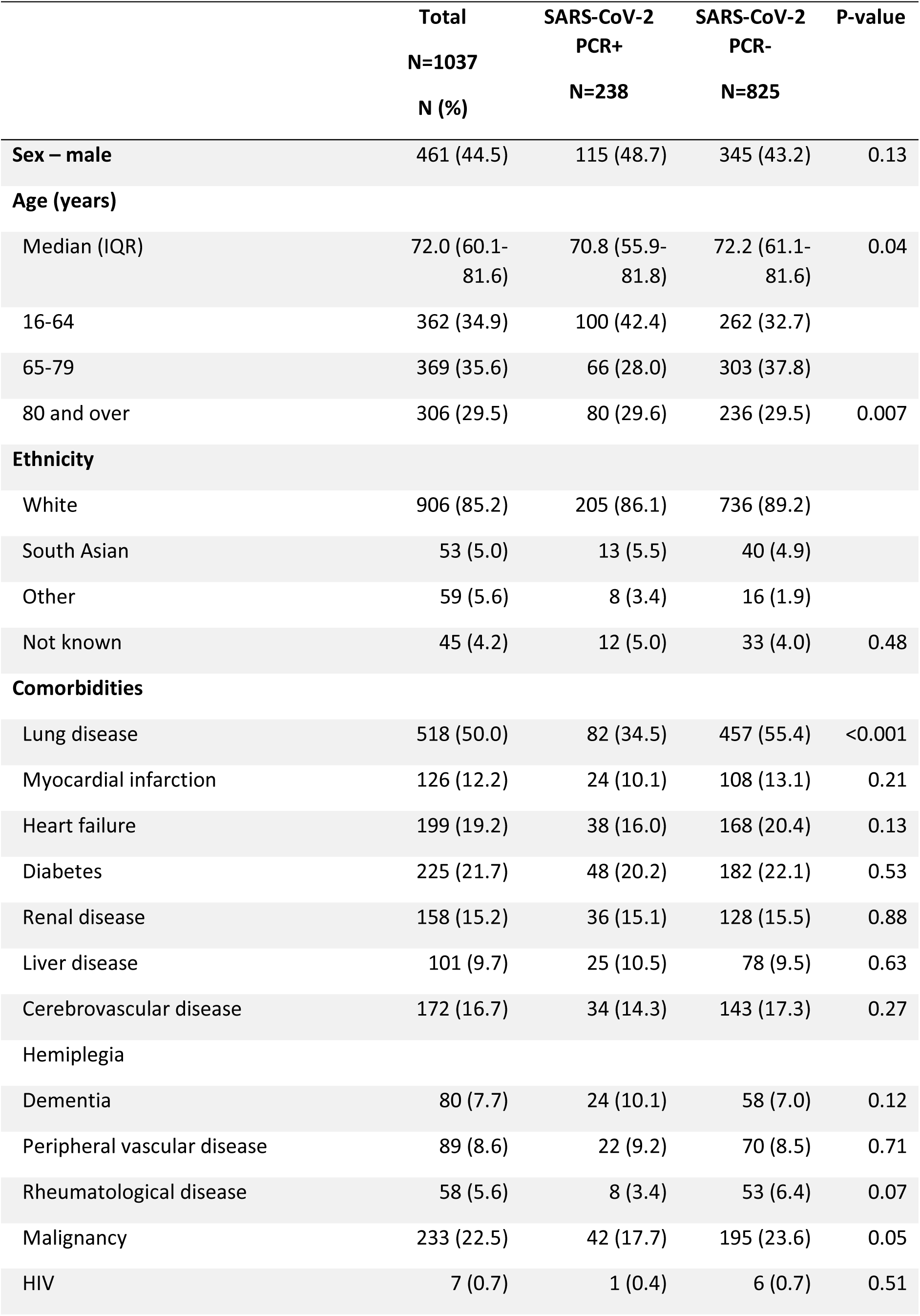
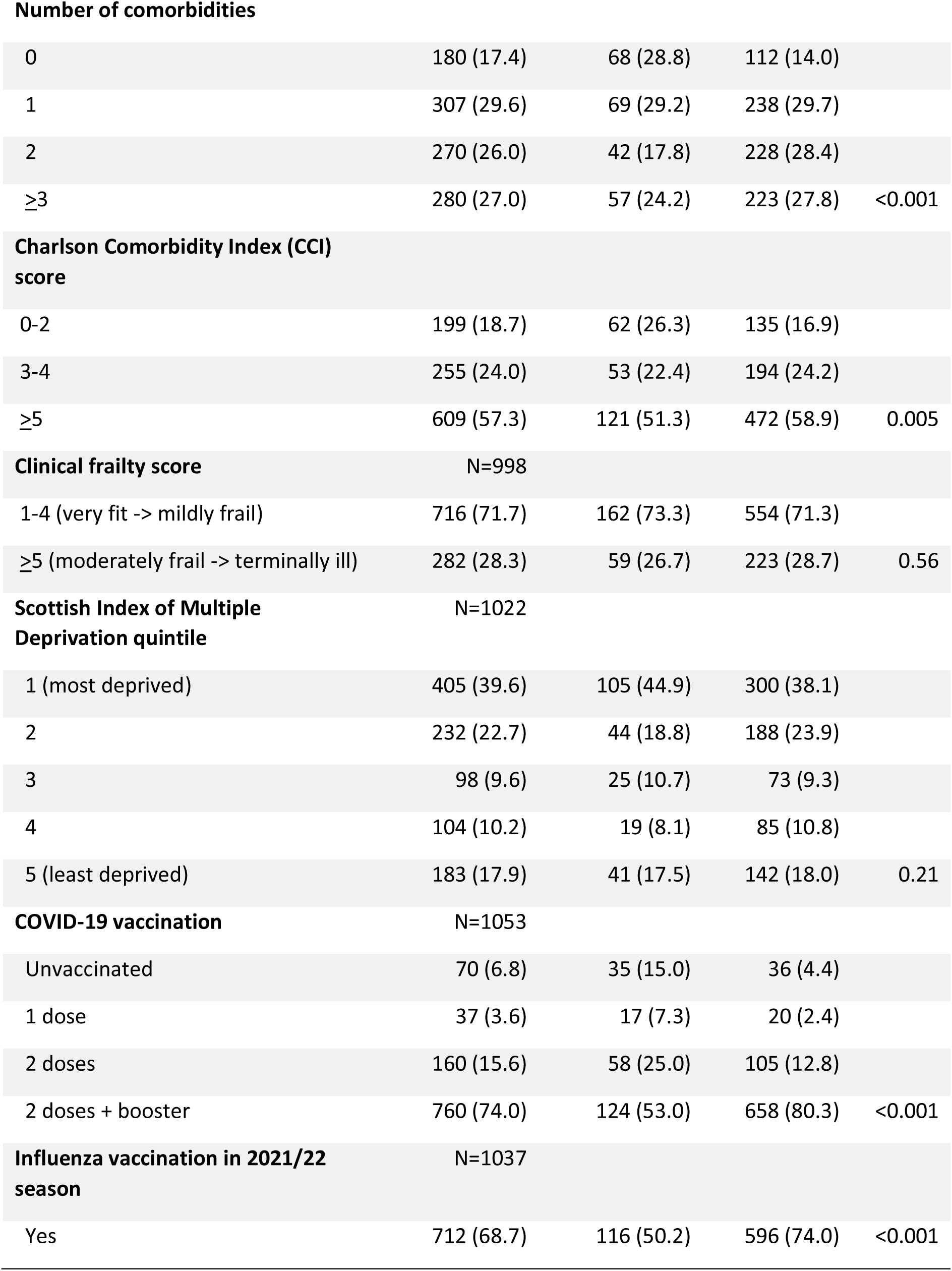
Demographic and clinical characteristics of recruited SARI cases, 23 November 2021 to 22 May 2022.

Scotland. Among 1027 individuals with available COVID-19 vaccination status, only 6.8% (n=70) were unvaccinated, while 74.0% (n=760) had received three doses. Over two-thirds (68.7%, n=712/1037) SARI cases had received influenza vaccination in the 2021/22 season.

### Virology, microbiology and radiology results

Overall, 236 (22.2%) SARI episodes were SARS-CoV-2 PCR positive, 33 (3.1%) were influenza A PCR positive, while fewer than five were influenza B PCR positive. Very few SARS-CoV-2 and influenza A co-infections were detected. Percentage PCR-positive for SARS-CoV-2 peaked in week 52 of 2021 (week beginning 27 December) at 52.1%, with a further peak in week 10 of 2022 (week beginning 7 March) at 38.8% (Figure 3A). These peaks in SARS-CoV-2 activity reflected national peaks in COVID-19-associated hospitalisations (Figure 3A).

SARS-CoV-2 PCR-positive SARI cases were slightly younger (median age 70.8 vs. 72.2 years, p=0.04), less likely to have underlying lung disease (34.5 vs. 55.4%, p<0.001) and had lower CCI score than SARS-CoV-2 PCR negative patients. Additionally, SARS-CoV-2 PCR positivity was highest in SARI cases who had never been vaccinated (49.3%, 35/71) and lowest in those who had received 3 vaccine doses (15.9%, 124/782).

Accompanying sequencing information was available for 86 (37.3%) of 236 SARS-CoV-2 PCR-positive SARI cases. Overall, 15 (17.4%) were Delta variants (B.1.617.2 and AY lineages), identified in COVID-19 cases in November and December 2022, while 29 (33.7%) and 31 (36.1%) were Omicron BA.1 and BA.2 subvariants, respectively (Figure 3A). BA.1 was detected in patients between the end of December 2021 and end of February 2022, while BA.2 was identified between February to end of May 2022. Sequences from 11 samples were unassigned.

Overall, 700 (65.9%) SARI episode had one or more microbiology samples sent; 143 had midstream specimen of urine (MSU) sent only, while 482 (45.3%) episodes had samples sent that were relevant to SARI presentation (lower respiratory culture (n=141), blood culture (n=378), or urinary Legionella antigen (n=104); Supplementary Figure 4). Among these, one or more significant microorganisms was identified in 61 (12.7%) episodes; more frequently in SARS-CoV-2 PCR-negative (14.0%, 54/386) than SARS-CoV-2 PCR-positive episodes (7.3%, 7/96). Among SARS-CoV-2 negative SARI episodes, the most frequently detected organisms were *Escherichia coli* (n=20; 14 from urinary source), *Haemophilus influenzae* (n=14) and *Streptococcus pneumoniae* (n=7). *Mycobacterial*, *Legionella* and *Aspergillus* species were also detected. In SARS-CoV-2 PCR-positive patients, *S. pneumoniae*, *Klebsiella pneumoniae* and several other organisms were detected. An additional 31 SARI episodes had a significant organism detected on MSU only, all but 8 episodes grew *E. coli*.

Chest radiology reports were identified in 987 (92.9%) SARI episodes. Consolidation was identified in similar proportion of SARS-CoV-2 PCR-positive and PCR-negative SARI episodes (39.1% vs. 39.2%, respectively). Of note, a higher proportion of SARI cases with consolidation had appropriate microbiology sent, compared to those without consolidation (53.9 vs. 39.8%, p<0.001).

### Clinical Presentation

The most commonly reported symptoms were shortness of breath (n=873, 79.3%), and cough (n=699; 65.8%), followed by fever (n=388, 36.5%). Gastrointestinal symptoms, such as vomiting (n=157, 14.8%) and diarrhoea (n=83, 7.8%) were also frequently reported. Only 21.7% (n=231) fulfilled WHO SARI case definition. The median duration of symptoms on presentation was 4 days (IQR 2-7).

Fatigue (35.6 vs. 24.3%), myalgia (8.5 vs. 3.4%), headache (10.2 vs. 4.6%), loss of smell (2.1 vs. 0.6%), loss of taste (2.5 vs. 0.6%), diarrhoea (11.9 vs. 6.7%), and altered conscious level (23.3 vs. 16.2%) were more frequently reported by SARS-CoV-2 PCR-positive SARI cases, while shortness of breath (82.3 vs. 66.5%) and purulent sputum (20.2 vs. 9.8%) were more common in SARS-CoV-2 PCR-negative cases (Supplementary Figure 2). Median duration of symptoms on presentation did not differ by SARS-CoV-2 PCR status (4 vs. 4 days).

### Management

Sixty percent of SARI episodes required supplementary oxygen (Table 2); most (53.9%, 553/1027) required ward-level oxygen only, 4.7% (49/1027) required high flow nasal oxygen (HFNO) or non-invasive ventilation, while 1.1% (11/1027) SARI episodes were mechanically ventilated. Maximum level of respiratory support did not differ by SARS-CoV-2 PCR status.

**Table 2.** Physiological parameters on admission, management and outcome of recruited SARI cases.

|  | Total<br>N (%) | SARS-CoV-2 PCR status |  | p-value |
| --- | --- | --- | --- | --- |
|  |  | Positive<br>N (%) | Negative<br>N (%) |  |
| Physiological parameters on admission |  |  |  |  |
| Temperature >38°c | 83/1010 (8.2) | 18/218 (8.3) | 65/792 (8.2) | 0.98 |
| Systolic blood pressure <90 mmHg | 26/1015 (2.6) | 6/219 (2.7) | 20/795 (2.5) | 0.86 |
| Diastolic blood pressure <60 mmHg | 153/1013 (15.1) | 37/219 (17.0) | 116/794 (14.6) | 0.40 |
| Heart rate >110 beats/minute | 246/1014 (24.3) | 58/219 (2.5) | 188/795 (23.7) | 0.39 |
| Oxygen saturation ≤92% on room air or require supplemental oxygen | 479/1012 (47.3) | 90/218 (41.3) | 389/794 (48.9) | 0.04 |
| Respiratory rate >30 breaths/minute | 115/1016 (11.3) | 28/219 (12.8) | 87/797 (10.9) | 0.44 |
| Glasgow coma scale <15 | 167/947 (17.6) | 44/207 (21.3) | 123/740 (16.6) | 0.12 |
| Chest X-Ray | N=987 | N=760 | N=227 |  |
| Evidence of consolidation | 386 (39.1) | 297 (39.1) | 89 (39.2) | 0.97 |
| Management |  |  |  |  |
| Antibiotics |  |  |  |  |
| On admission | 600/1019 (58.9) | 96/219 (43.8) | 504/800 (63.0) | <0.001 |
| During inpatient stay | 794/1063 (74.7) | 126/236 (53.4) | 668/827 (80.8) | <0.001 |
| Antivirals |  |  |  |  |
| On admission | 16/1019 (1.6) | <5/219 (1.8) | 12/800 (1.5) | 0.73 |
| During inpatient stay | 69/1063 (6.5) | 26/236 (11.0) | 43/827 (5.2) | 0.001 |
| Steroids |  |  |  |  |
| On admission | 327/1019 (32.1) | 84/219 (38.4) | 243/800 (30.4) | 0.03 |
| During inpatient stay | 457 (43.0) | 121/236 (51.3) | 336/827 (40.6) | 0.004 |
| Maximum level of care | N=1018 | N=218 | N=800 |  |
| Ward | 936 (91.9) | 192 (88.1) | 744 (93.0) |  |
| High dependency Unit | 69 (6.8) | 22 (10.1) | 47 (5.9) |  |
| Intensive Care Unit | 13 (1.3) | <5 (1.8) | 9 (1.1) | 0.06 |

| **Maximum level of respiratory support** | **N=1027** | **N=223** | **N=804** |  |
| --- | --- | --- | --- | --- |
| No supplemental oxygen | 414 (40.3) | 96 (43.1) | 318 (39.6) |  |
| Ward-level oxygen | 553 (53.9) | 116 (52.0) | 437 (54.4) |  |
| HFNO/NIV | 49 (4.7) | 8 (3.6) | 41 (5.0) |  |
| Invasive mechanical ventilation | 11 (1.1) | <5 (1.3) | 8 (1.0) | 0.63 |
| **Outcome** | **N=1061** | **N=235** | **N=826** |  |
| Discharged home | 956 (90.1) | 208 (88.5) | 748 (90.6) |  |
| Transferred to another health facility or palliative discharge | 22 (2.1) | 7 (3.0) | 15 (1.8) | 0.23 |
| Died in hospital | 83 (7.8) | 20 (8.5) | 63 (7.7) | 0.68 |
| Death within 30-days of admission | 107 (10.1) | 27 (11.5) | 80 (9.7) | 0.41 |
| Length of hospital stay* - median (IQR) | 7 (4-13) | 7 (4-13) | 7 (4-12) | 0.95 |

PCR, polymerase chain reaction; HFNO, high flow nasal oxygen; NIV, non-invasive ventilation. As per conventions for statistical disclosure control, the threshold for privacy risk of 'n' observations have been set to 5. Any values <5 has been ascribed '<5'.
\*In patients who survived to hospital discharge.

In terms of maximum level of care, 8.1% (n=82/1018) required admission to critical care. A slightly higher proportion of SARS-CoV-2 PCR-positive SARI episodes required admission to HDU (10.0% vs. 5.9%) and ICU (1.8% vs. 1.1%), compared to SARS-CoV-2 PCR negative SARI episodes.

Antimicrobial use was high, with 58.9% (600/1019) and 74.7% (794/1063) receiving antibiotics on admission and during their admission, respectively (Table 2). Just over half of SARI episodes (54.5%, 327/600) who were prescribed empirical antibiotics had a respiratory or blood culture sent, compared to 31.0% (130/457) of those who did not receive empirical antibiotics. Inpatient antimicrobial prescription was higher among SARI episodes that tested negative for SARS-CoV-2 or influenza viruses (80.9% vs. 53.4%, p<0.001), had one or more significant microbiological organisms detected (89.1 vs. 73.3%, p<0.001), and among SARI episodes that involved admission to critical care (86.6% vs. 75.8%, p=0.03) With regards to antiviral treatment, 42.4% of SARI episodes associated with a positive influenza A or B PCR were prescribed oseltamivir, including the patients with influenza A and SARS-CoV-2 co-infection (Supplementary Table 1). Among SARS-CoV-2 PCR-positive SARI episodes (n=238), two (0.8%) received remdesivir, while around a third (35.2%, n=84) were prescribed steroids.

### Outcome

At the time of final data extraction, patient outcome was available for all but two SARI episodes. Most were discharged home (90.1%, n=956/1061), 7.8% (n=83) died in hospital, while 28-day mortality was 10.1% (n=107) (Table 2). Among patients who were alive at discharge, median length of hospital stay was 6 days (IQR 3-12 days). Neither duration of hospital admission, all-cause in-hospital, nor 28-day mortality differed by SARS-CoV-2 status.

### Factors associated with in-hospital mortality

On univariable analysis, male sex, older age group, higher CCI score, higher frailty score and less deprived SIMD quintiles were associated with in-hospital death, as were low diastolic blood pressure (<60mmHg), tachycardia (heart rate >110 beats/minute), tachypnoea (respiratory rate >30 breaths/minute), GCS <15 and consolidation on chest X-ray. Conversely, influenza and SARS-CoV-2 vaccination status, in addition to SARS-CoV-2 PCR positivity had no impact on mortality (Table 3).

**Table 3.**
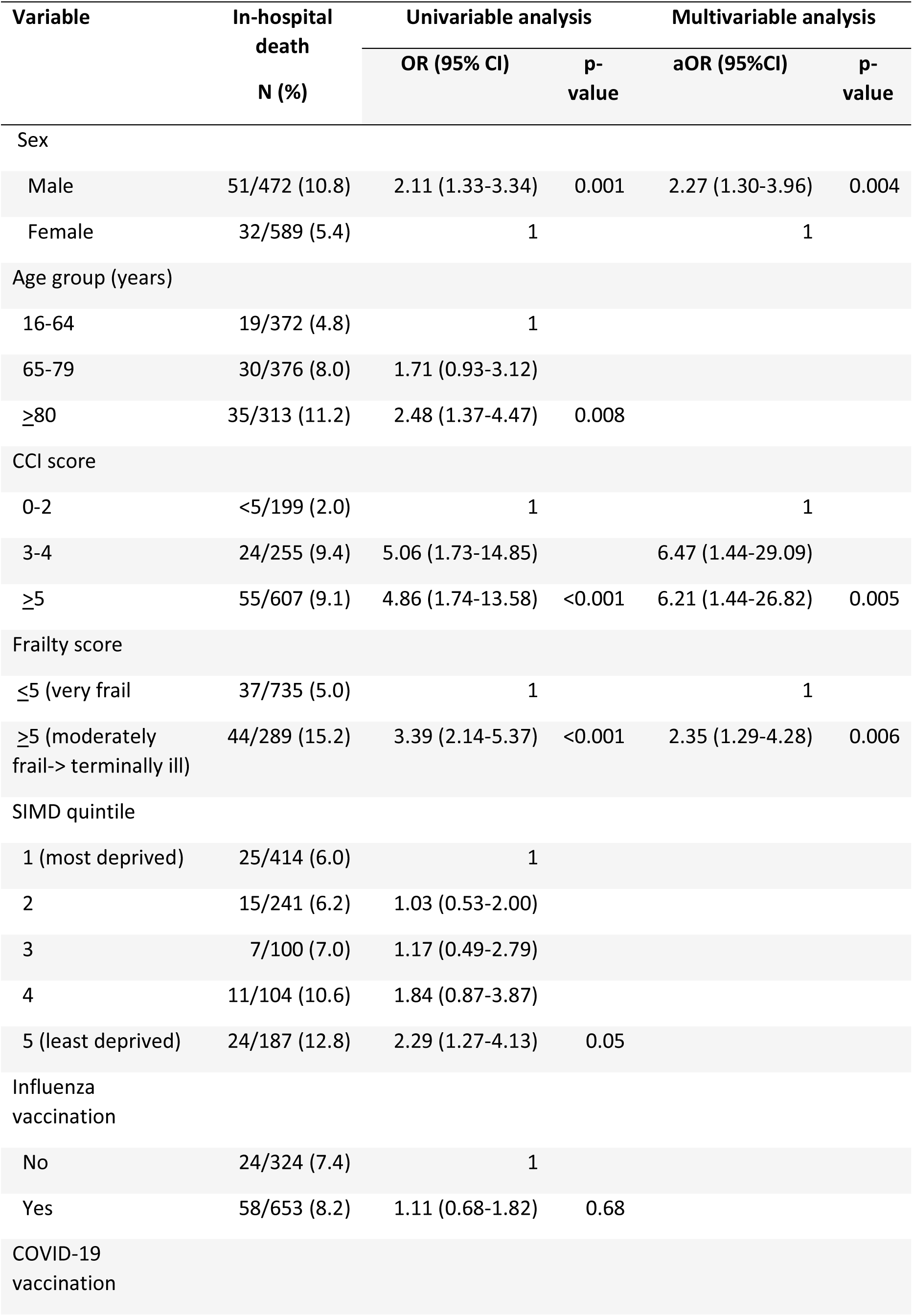
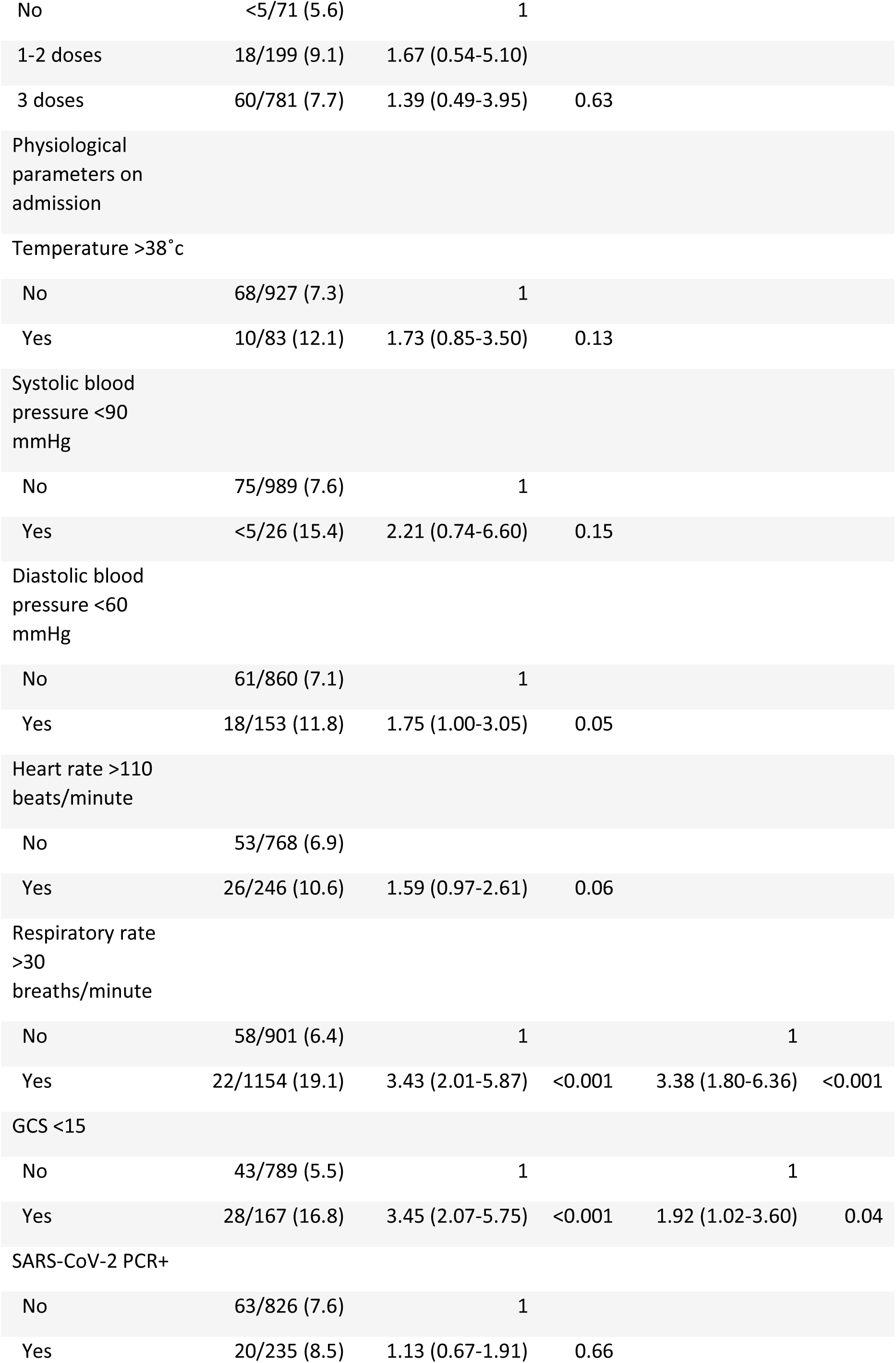
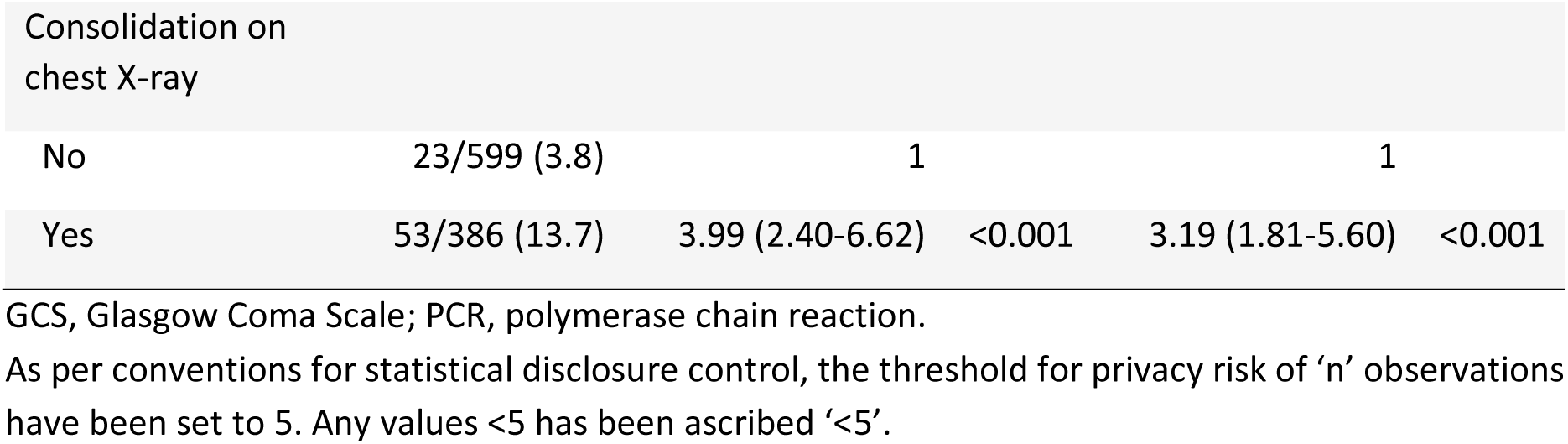
Factors associated with in-hospital mortality among recruited SARI cases.

On multivariable analysis, male sex (adjusted OR (aOR) 2.27, 95%CI 1.30-3.96), increasing CCI score (3-4: aOR 6.47, 95%CI 1.44-29.09; ≥5: aOR 6.21, 95%CI 1.44-26.82; vs. 0-2), frailty score ≥5 (aOR 2.35, 95%CI 1.29-4.28, vs <5) respiratory rate >30 on admission (aOR 3.38, 95%CI 1.80-6.36), GCS <15 on admission (aOR 1.92, 95%CI 1.02-3.60) and chest X-ray consolidation (aOR 3.19, 95%CI 1.81-5.60) were independent predictors of in-hospital mortality among SARI cases.

### Prediction of in-hospital mortality by 4C mortality score and CURB-65

Of 193 patients with a positive SARS-CoV-2 PCR in whom 4C mortality score could be calculated, 16.6% (n=32), 24.3% (n=47), 54.9% (n=106) and 4.2% (n=8) were in the low, intermediate, high and very high risk groups, respectively (Supplementary Table 2). In-hospital mortality was 0%, 4.2%, 11.1% an 25.0% in the respective groups. 4C score showed high discrimination for mortality (AUROC 0.796, 95%CI 0.693-0.898; Supplementary Figure 5A). Overall observed vs. predicted probability of in-hospital mortality were equal (8.3%, CITL=0), and calibration was excellent across the range of risks (slope=1, Brier score 0.066; Supplementary Figure 6A). 4C score also demonstrated reasonable discrimination and calibration among SARS-CoV-2 PCR-negative patients (AUROC 0.740, 95% CI 0.676-0.804; CITL=0; slope=1; Brier score 0.062; Supplementary Table 2; Supplementary Figure 5B & 6B). CURB-65 demonstrated good calibration among SARS-CoV-2 PCR-positive and negative SARI cases, but discrimination was lower than 4C score in both groups (AUROC SARS-CoV-2 PCR+: 0.751, 95%CI 0.634-0.867; SARS-CoV-2 PCR-: 0.714, 95%CI 0.650-0.777; Supplementary Figure 5C & D, 6C & D).

### Discharge diagnosis

ICD10-coded discharge diagnoses were available for 1005 (94.5%) SARI cases. The most common diagnoses were community-acquired pneumonia (CAP)/ lower respiratory tract infection (LRTI; n=392, 39.0%), exacerbation of chronic obstructive pulmonary disease (COPD) (n=201) and COVID-19 (n=200, 19.9%) (Supplementary Table 3). Of note, COVID-19 was not included as a discharge diagnosis in 16.7% (37/221) SARI cases with a positive SARS-CoV-2 PCR.

In terms of combinations of diagnoses, CAP/LRTI and other diagnoses was the most frequent combination (n=113), followed by CAP/LRTI (n=71), COPD exacerbation (n=60), COVID-19 and other diagnosis (n=60) as well as COVID-19 and CAP/LRTI (n=40) (Supplementary Figure 7).

## Discussion

We successfully implemented an enhanced syndromic surveillance by integrating routinely collected data with additional research resource, providing PHS and clinicians near real-time snapshots of the contribution of influenza viruses and SARS-CoV-2 to hospitalised SARI. Proportions of influenza and SARS-CoV-2 PCR positivity were reflective of national hospitalisation figures. SARS-CoV-2 was the dominant pathogen among SARI cases, with two waves between November 2021 and May 2022 driven by the emergence of Omicron BA.1 and BA.2 variants, respectively. In contrast, influenza activity was low, with a small wave of influenza A in March 2022. Around 60% required supplemental oxygen, but very few (∼1%) required mechanical ventilation. In-hospital mortality was ∼8%; male sex, multimorbidity, frailty, tachypnoea, altered mental state and radiological consolidation predicted mortality. These data supported both in-hospital and PHS decision making.

During the first two years of the pandemic, numerous studies provided detailed description of the clinical epidemiology of hospitalised patients with COVID-19.^8^ ^9^ In contrast, the evolving epidemiology of COVID-19 alongside the re-emergence of influenza infections in hospitalised patients as we transitioned out of the pandemic phase has been poorly characterised. Two European studies reported on their newly established SARI surveillance in the 2021/22 season, both part of the European network, E-SARI-NET. Using routine ED records, Brady *et al*. identified a high prevalence of SARS-CoV-2 infection (52% of those tested), and low positivity for influenza (4.3%) and RSV (2.3%) among SARI cases in an Irish hospital,^10^ while Torres *et al*. identified SARI cases in two Portugese regions using electronic health registries,^11^ correlating patients with ICD-10 codes for infuenza-like illness, cardiovascular diagnosis, respiratory diagnosis, and respiratory infection in their primary admission diagnosis with weekly influenza and COVID-19 incidence. Several studies compared mortality between COVID-19 and influenza in hospitalised patients in 2021/22^12^ ^13^ and 2022/23^14^; all found a higher in-hospital (7 vs. 4.4%)^13^ or 30-day mortality (6-8% vs. 2.5-3.8%)^12^ ^14^ in patients infected with with SARS-CoV-2 Omicron variants compared to influenza viruses, particularly among unvaccinated individuals.

The symptom profile of COVID-19 has evolved with each new variant, alongside increasing population immunity from both infection and vaccination.^15^ Since characterising the changing clinical presentation of COVID-19 was an objective of the study, we opted for a less stringent SARI case definition than the WHO definition (reported or measured fever of ≥38°C and cough; onset within the last 10 days; and requiring hospitalisation)^16^ to enhance sensitivity. The requirement for fever tends to exclude older patients, since fever is often attenuated or absent in this group.^17^ Indeed, 45% of SARI cases in our study aged <65 years had a reported or recorded fever, compared to 35% and 33% of those aged 65-79 and ≥80 years, respectively. Reflective of this, just over 1 in 5 recruited SARI cases fulfilled the WHO SARI criteria.

We identified different symptom profiles between SARS-CoV-2 PCR-positive and PCR-negative SARI cases; influenza-like symptoms, such as fatigue, myalgia, headache, in addition to loss of smell and taste were more frequently reported by predominantly Omicron-infected COVID-19 cases, while shortness of breath and purulent sputum were more commonly reported by SARS-CoV-2 negative SARI cases. There were insufficient number of influenza PCR-positive cases to evaluate their symptom profile separately, or the impact of influenza SARS-CoV-2 co-infections.

Although influenza activity was low compared to SARS-CoV-2 in the study period, there was similar probability of a SARI case being infected with influenza and SARS-CoV-2 in March 2022 (Figure 3). The availability of these data in near real-time allowed us to rapidly alert ED and acute medicine physicians, ensuring that both viruses were considered in the differential diagnosis.

Overall in-hospital mortality was 7.8%; mortality increased with age, but were equivalent by SARS-CoV-2 status. Male sex, multimorbidity (high CCI score), frailty, tachypnoea, altered mental state, and consolidation on chest X-ray predicted in-hospital mortality. In particular, those with a CCI score >2 had a 6 fold-increased odds of death compared to those with a CCI score 0-2. In the univariable analysis, SARI cases from less deprived quintiles were associated with higher mortality. This was likely confounded by age and comoribidites (i.e. those from less deprived backgrounds were older), hence this association disappeared in the multivariable analysis.

COVID-19 morbidity and mortality were substantially lower than reported in hospitalised patients in previous waves.^8^ ^18^ The majority of SARI cases (>70%) were infected with Omicron subvariants, which were associated with lower risk of ICU admission and death, compared to Delta.^19^ Furthermore, three-quarters of recruited SARI cases had received three vaccine doses. Nevertheless, 4C mortality score continued to demonstrate good discrimination for in-hospital mortality in a highly vaccinated patient cohort infected by Delta and Omicron variants, as demonstrated by other studies.^20^ ^21^ Interestingly, 4C score also showed reasonable predictive performance in non-COVID-19 SARI cases (AUROC 0.740). In comparison, CURB-65 demonstrated poorer discrimination in both groups. Further assessment of 4C score in hospitalised SARI cohorts is warranted to determine whether it could be deployed as a risk stratification tool in the context of non-COVID-19 SARI.

A significant microorganism was identified in 8.7% of the SARI cohort, though fewer than half of recruited SARI cases had appropriate microbiological samples sent. Conversely, antimicrobial prescription was high; although lower in COVID-19 patients, nearly 60% of SARI episodes received empirical antimicrobials. Low rate of microbiological testing has been described in pneumonia cohorts,^22^ ^23^ and hospitalised COVID-19 patients,^24^ likely due to a combination of clinical pressure in acute care areas, and low sensitivity of current micriobiological diagnostics. Nevertheless, given the high prevalence of SARI presentations and concerns about emerging antimicrobial resistance, microbiological testing is critical, not only to ensure appropriate antimicrobial treatment, but to generate up-to-date epidemiological data. Bacterial coinfections are prevalent in hospitalised patients with influenza, but less frequent in COVID-19.^24^ Ongoing evaluation of more sensitive molecular diagnostics as well as biomarkers predictive of a bacterial aetiology are needed to support clinician decisions regarding antimicrobial prescriptions.

We encountered several challenges with the study. First, although data collection was primarily undertaken by the research team, we aimed to engage treating clinicians to identify eligible patients, and record data on patient demographics, frailty score and presenting symptoms, as these details would be ascertained during patient assessment. However, only ∼5% SARI cases were inputted by non-research staff. A subsequent qualitative study found that ED clinical staff recognised the importance of research to improve patient care, but cited time constraints and concerns about the impact on clinical duties as barriers to study participation, highlighting that integration of SARI surveillance in a clinical setting without dedicated personnel would be extremely difficult.^25^ Similar challenges were encountered by Irish SARI surveillance.^10^ Second, we were only able to identify residual respiratory samples from 15% of recruited SARI cases, hence were unable to comprehensively assess for infections by other respiratory viruses. We have switched to a consented study for 2022/23 and 2023/24 recruitment to ensure an additional respiratory sample can be obtained.

The key limitation of this study was that it involved a single site, thus findings may not be generalisable nationally. Nevertheless, it is the largest hospital in Scotland that covers a population of broad socioeconomic background. Small numbers of influenza PCR-positive SARI cases meant this group could not be evaluated separately. We were also unable to study the impact of influenza and SARS-CoV-2 co-infections.

In summary, we successfully implemented of an enhanced SARI surveillance, leveraging linkage of routinely available clinical data to limited research data collection, to provide near real-time data on the contribution of SARS-CoV-2 and influenza among hospitalised SARI patients, in addition to detailed characterisation of symptom profile, management, illness severity and outcome. These data are being used to optimise SARI management pathways; we envisage the reintroduction of care bundles for SARI, with particular emphasis on improving microbiological testing and developing guidance to support clinicians’ decision making regarding antimicrobial and antiviral prescription. Lastly, the recent re-emergence of measles, *Bordetella pertussis*, *Mycoplasma pneumoniae,* and zoonotic influenza spillover events^26^ ^27^ underscore the importance of pathogen-agnostic surveillance with clinical metadata to identify emerging and re-emerging respiratory pathogens. We recommend expansion of this enhanced surveillance to additional UK sites, including paediatric units.

## Data Availability

All data produced in the present study are available upon reasonable request to the authors

## Acknowledgements

We would like to acknowledge National Education Scotland, NHS Greater Glasgow and Clyde Safehaven and e-Health, in addition to the Clinical Research Facility ED including the ED Research nurse team supporting the inception and delivery of this study.

## Funding

This work was supported by Public Health Scotland. The funder had no involvement in the study design, collection, analysis and interpretation of data, writing of the report, or in the decision to submit this article for publication.

## Author contributions

Conceptualisation: AH and DJL. Formal analysis: AH, MM. Investigation: all authors. Data curation: AH. Visualization: AH. Writing – original draft: AH. Writing – review & editing: Supervision – AH and DJL. Funding acquisition: AH and DJL.

## Notes

### Competing Interest Statement

The authors have declared no competing interest.

### Funding Statement

This study was funded by Public Health Scotland.

### Author Declarations

Ethics approval was granted by the North of Scotland Research Ethics Committee (ref. 21/NS/1036) and the National Health Service Greater Glasgow and Clyde Biorepository (ref. 16/WS/0207)

