## Supplementary Materials for "Adults with Severe Acute Respiratory Illness in Glasgow, 2021-22: A Prospective Cohort Study"

### **Supplementary Tables**

#### **Supplementary Table 1.** Antimicrobial, antiviral and steroid prescription on admission and during inpatient stay

|  | On admission  N (%) | During admission  N(%) |
| --- | --- | --- |
| Antivirals | 16/1019 (1.6) | 69 (6.5) |
| Oseltamivir | 14 (1.6) | 26 (2.4) |
| Remdesivir | <5 (0.2) | 21 (2.0) |
| Aciclovir | 10 (0.9) | 10 (0.9) |
| Antimicrobials | 600/1019 (58.9) | 794/1063 (74.7) |
| Amoxicillin | 36 (35.4) | 430 (40.5) |
| Clarithromycin | 147 (14.4) | 180 (16.9) |
| Doxycycline | 94 (9.2) | 231 (21.7) |
| Co-amoxiclav | 61 (6.0) | 140 (13.2) |
| Gentamicin | 45 (4.4) | 82 (7.7) |
| Levofloxacin | 28 (2.7) | 39 (3.7) |
| Piperacillin-tazobactam | 21 (2.1) | 59 (5.6) |
| Co-trimoxazole | 7 (0.7) | 42 (4.0) |
| Ciprofloxacin | - | 22 (2.1) |
| Ceftriaxone | - | 14 (1.3) |
| Temocillin | - | 5 (0.5) |
| Steroids | 327/ 1019 (32.1) | 457/1063 (43.0) |
| Dexamethasone | 92 (9.0) | 135 (12.7) |
| Hydrocortisone | 53 (5.2) | 34 (3.2) |
| Prednisolone | 184 (18.1) | 328 (30.9) |

As per conventions for statistical disclosure control, the threshold for privacy risk of ‘n’ observations have been set to 5. Any values <5 has been ascribed ‘<5’.

#### **Supplementary Table 2.** In-hospital mortality in SARI cases by 4C mortality score risk category and SARS-CoV-2 PCR status

| Risk group | 4C  Mortality  Score | All | | SARS-CoV-2 PCR+  N=193 | | SARS-CoV-2 PCR-  N=701 | |
| --- | --- | --- | --- | --- | --- | --- | --- |
|  |  | **N Patients (%)** | **Mortality (%)** | **N Patients (%)** | **Mortality (%)** | **N Patients (%)** | **Mortality (%)** |
| Low | 0-3 | 88 (9.8) | 0 (0) | 32 (16.6) | 0 (0) | 56 (8.0) | 0 (0) |
| Intermediate | 4-8 | 245 (27.4) | 6 (2.5) | 47 (24.3) | <5 (4.2) | 198 (28.3) | 4 (2.0) |
| High | 9-14 | 522 (58.4) | 47 (8.8) | 106 (54.9) | 12 (11.1) | 416 (59.3) | 35 (8.4) |
| Very high | ≥15 | 39 (4.4) | 13 (33.3) | 8 (4.2) | <5 (25.0) | 31 (4.4) | 11 (35.5) |

PCR, polymerase chain reaction

#### **Supplementary Table 3.** Discharge diagnoses based on ICD-10 code

|  | N=1005 (%) |
| --- | --- |
| COVID-19 | 200 (19.9) |
| Influenza infection | 27 (2.7) |
| Pneumonia/ lower respiratory tract infection | 392 (39.0) |
| Exacerbation of COPD | 201 (10.0) |
| Other respiratory infection | 11 (1.1) |
| Aspiration pneumonia | 15 (1.5) |
| Pulmonary embolism | 35 (3.5) |
| Other respiratory condition | 128 (12.7) |
| Lung malignancy | 26 (2.6) |
| Myocardial infarction | 9 (0.9) |
| Cardiac failure | 794 (74.7) |
| Other cardiac condition | 92 (9.2) |
| Other infection | 140 (13.9) |
| Other diagnosis | 429 (42.7) |

COVID-19, coronavirus disease 2019; COPD, chronic obstructive pulmonary disease.

### **Supplementary Figures**

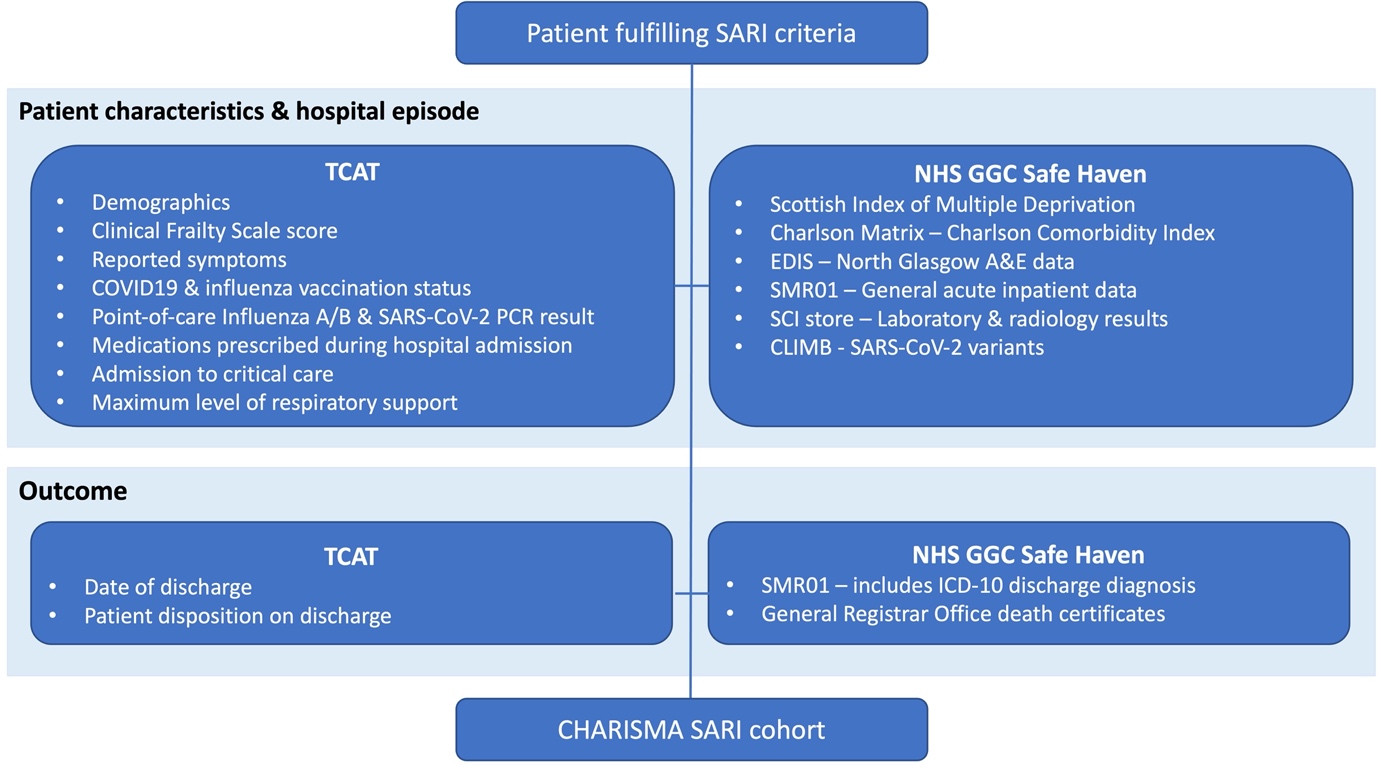

#### **Supplementary Figure 1**. Data linkage diagram

TCAT, Turas Clinical Assessment Tool; PCR, polymerase chain reaction; GGC, Greater Glasgow and Clyde; EDIS, Emergency Department Information System; CLIMB, Cloud Infrastructure for Microbial Bioinformatics; ICD-10, International Classification of Diseases, Tenth Revision; SARI, severe acute respiratory illness

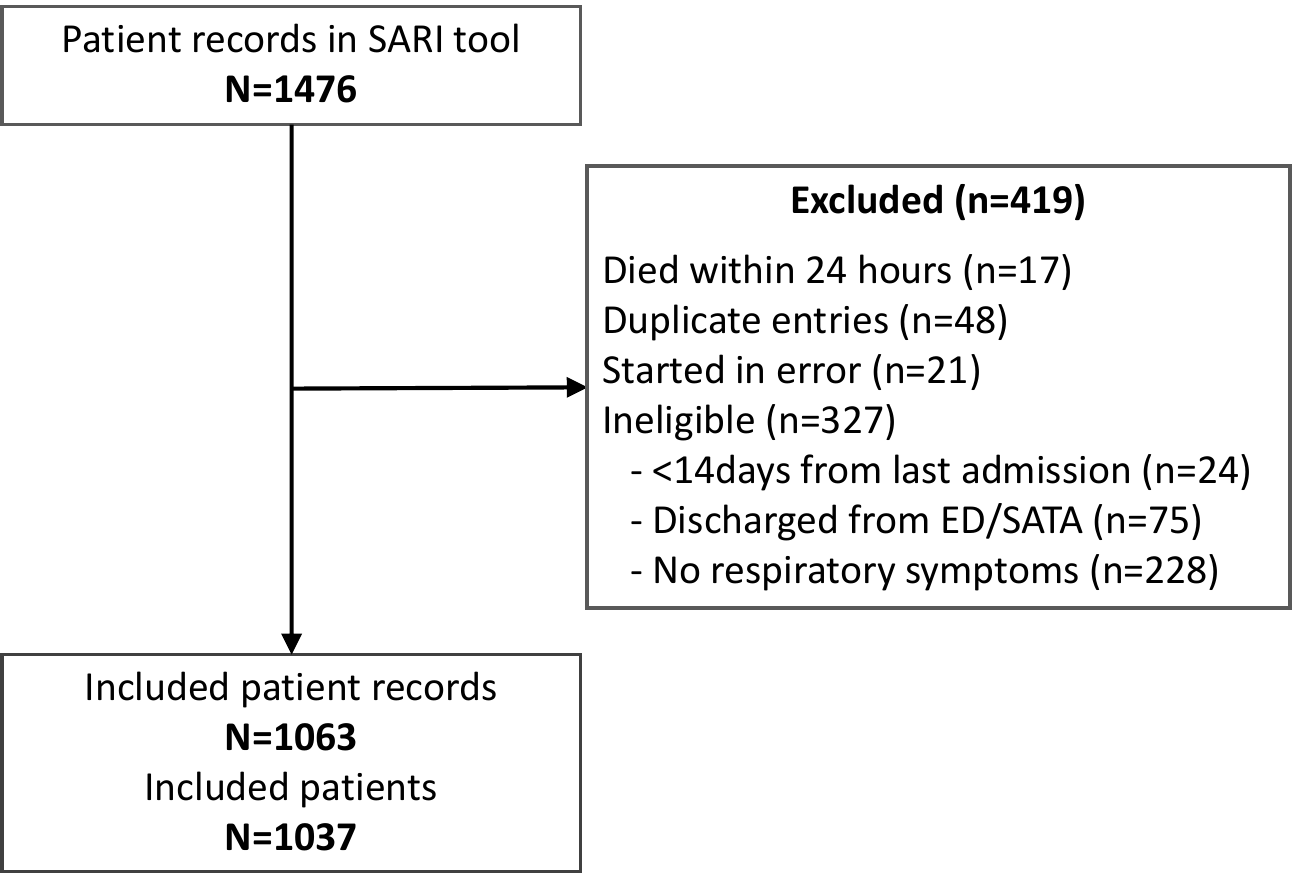

#### **Supplementary Figure 2.** Flow chart of patient screening and recruitment

SARI, severe acute respiratory illness; ED, Emergency Department; SATA, Specialist Assessment and Treatment Area

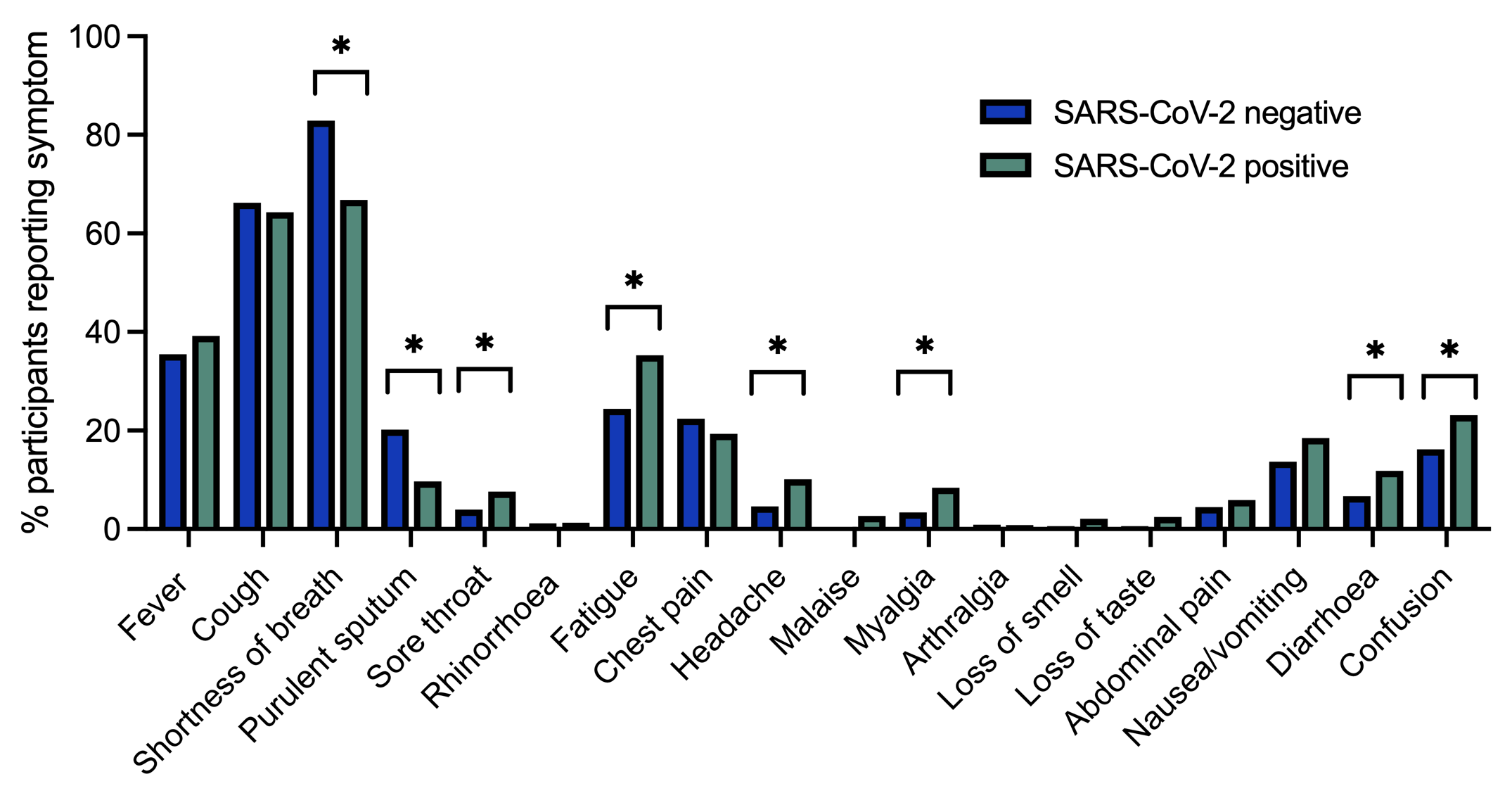

#### **Supplementary Figure 3.** Reported symptoms by SARS-CoV-2 PCR status

SOB, shortness of breath; PCR, polymerase chain reaction.

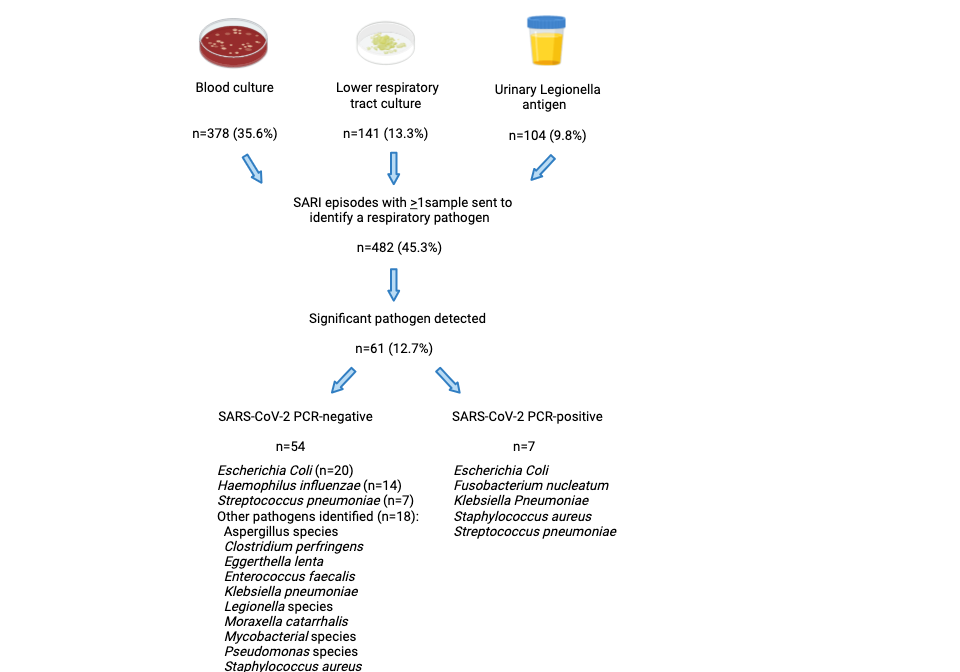

#### **Supplementary Figure 4.** Microbiological investigations and significant bacterial and fungal pathogens detected

| 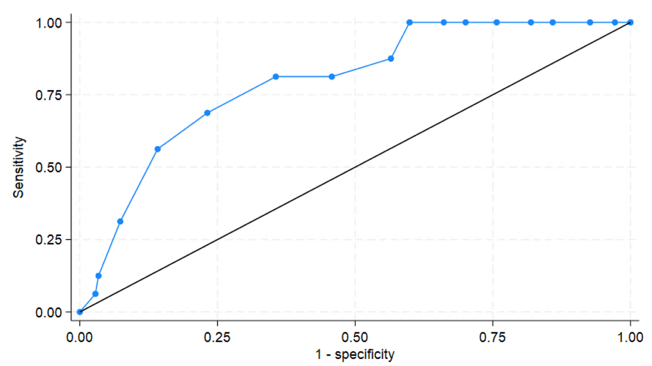  **A)** | 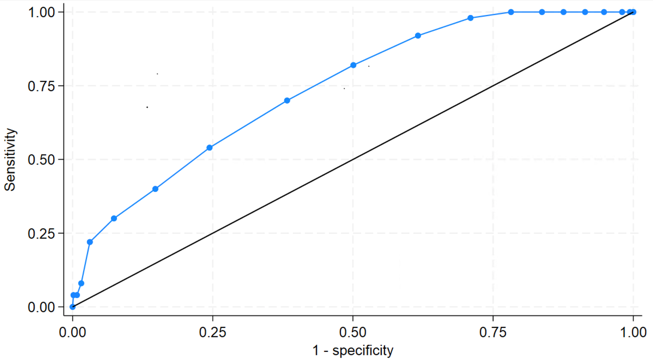  **B)** |
| --- | --- |
| 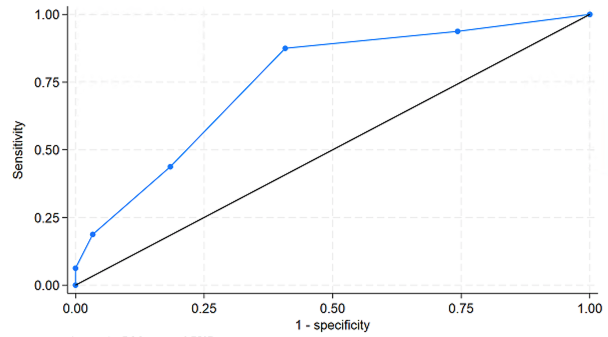  **C)** | 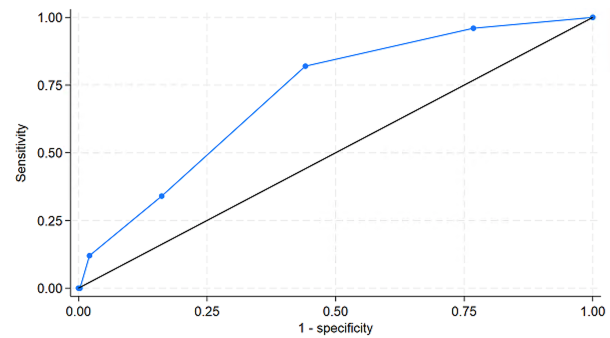  **D)** |

**Supplementary Figure 5.** Predictive performance of 4C mortality and CURB-65 score in severe acute respiratory illness (SARI) cases. A) 4C mortality score in SARS-CoV-2 PCR-positive severe acute respiratory illness (SARI) cases (n=163); area under the receiver operating characteristic (AUROC) 0.796, 95%CI 0.693-0.898) and B) SARS-CoV-2 PCR-negative SARI cases (n=701, AUROC 0.740, 95%CI 0.676-0.804); **C)** CURB-65 in SARS-CoV-2 PCR-positive SARI cases (n=195, AUROC 0.751, 95%CI 0.634-0.867) and **D)** SARS-CoV-2 PCR-negative SARI cases (n=705; AUROC 0.714, 95%CI 0.650-0.777)

| 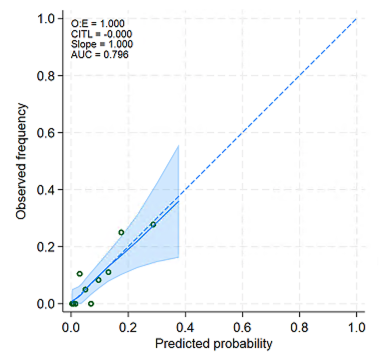  **A)** | 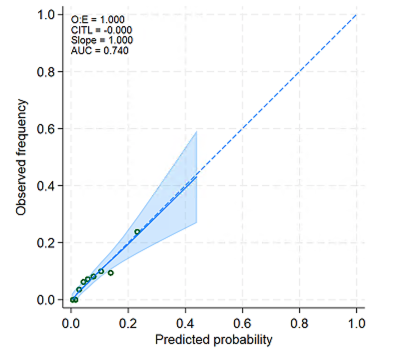  **B)** |
| --- | --- |
| 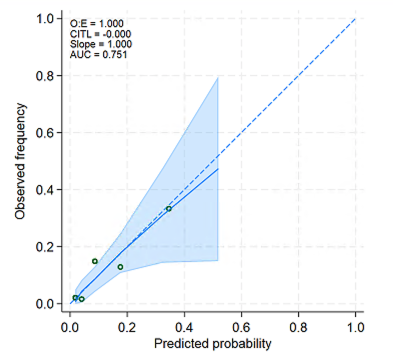  **C)** | 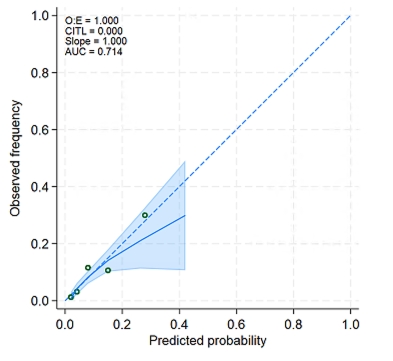  **D)** |

**Supplementary Figure 6.** Predicted versus observed probability of in-hospital mortality in A) SARS-CoV-2 PCR positive SARI cases using 4C mortality score; B) SARS-CoV-2 PCR negative SARI cases using 4C mortality score; C) SARS-CoV-2 PCR positive SARI cases using CURB-65 score; D) SARS-CoV-2 PCR negative SARI cases using CURB-65 score.

O:E, observed: expected; CITL, calibration in-the-large; AUC, area under the receiver operating characteristic.

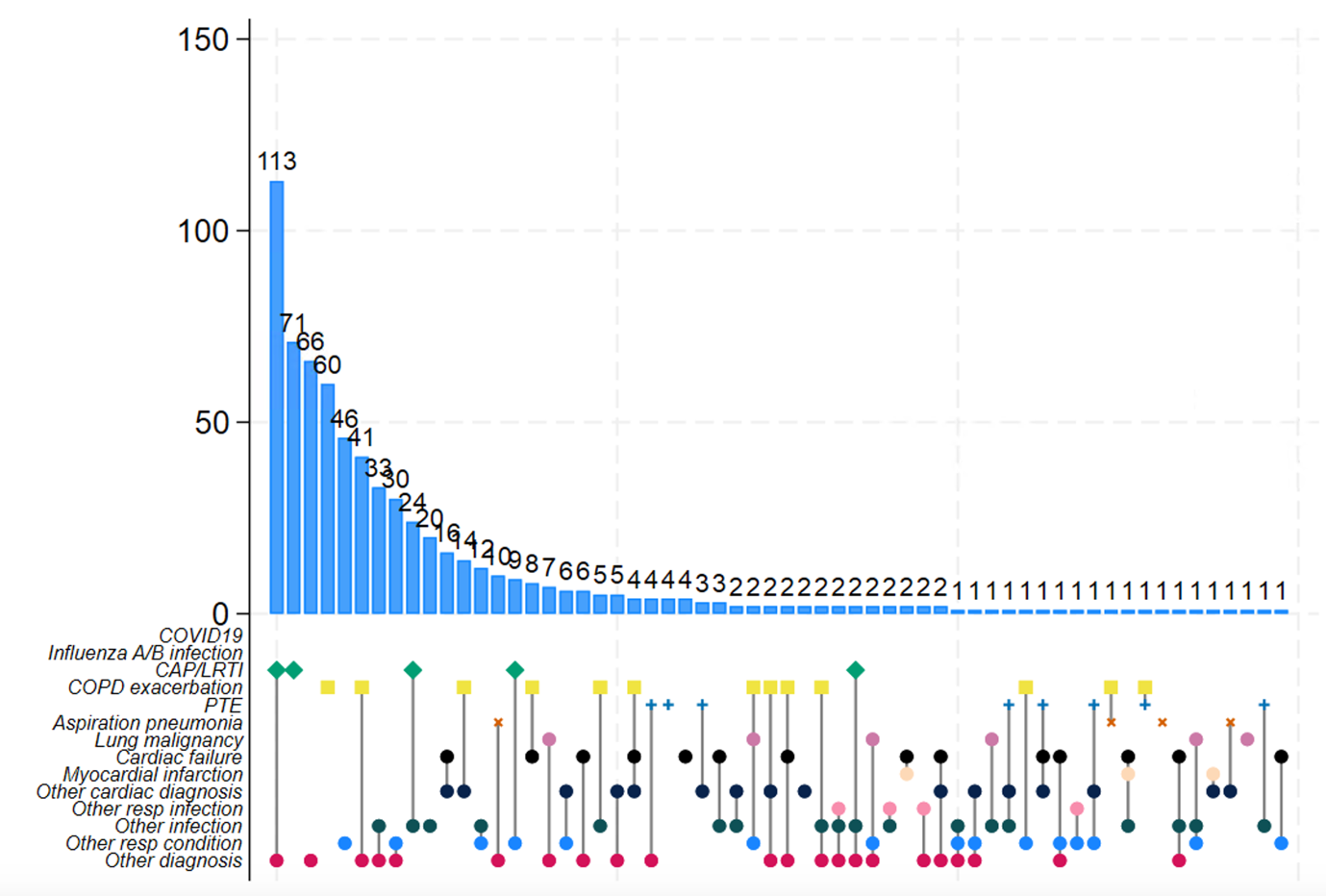

**Supplementary Figure 7.** Upset plot of ICD-10 coded discharge diagnoses of recruited SARI cases

COVID-19, coronavirus disease 19; CAP, community-acquired pneumonia; LRTI, lower respiratory tract infection; COPD, chronic obstructive pulmonary disease; PTE, pulmonary thromboembolism.
